# Unraveling the Myths of *R*_0_ in Controlling the Dynamics of COVID-19 Outbreak: a Modelling Perspective

**DOI:** 10.1101/2020.04.25.20079319

**Authors:** Mohd Hafiz Mohd, Fatima Sulayman

## Abstract

COVID-19 is an emerging and rapidly evolving pandemic around the world, which causes severe acute respiratory syndrome and results in substantial morbidity and mortality. To examine the transmission dynamics of COVID19 and its interactions with some exogenous factors such as limited medical resources and false detection problems, we employ a simple epidemiological model and analyse this system using modelling and dynamical systems techniques. We discover some contrasting findings with respect to the observations of basic reproduction number, and we investigate how the issues of limited medical resources and false detection problems affect the COVID-19 pandemic outbreak.

## 1. INTRODUCTION

Recently, a novel coronavirus called SARS-CoV-2 (or previously referred to as 2019-nCoV) has been identified as the causes of a severe acute respiratory syndrome and a cluster of pneumonia cases in Wuhan, China^1, 2^. This virus rapidly spreads throughout China, followed by increasing number of cases across the globe, which lead to pandemic outbreaks in different countries such as United States, Italy, Malaysia^2^. The World Health Organization terms the disease associated with SARS-CoV-2 virus as COVID-19, which refers to coronavirus disease 2019.

In curbing the COVID-19 pandemic in Malaysia, the government through its Ministry of Health has announced a targeted testing approach so as to optimally use the country’s limited medical resources^3^. In targeted testing, the tests are conducted based on location of COVID-19 hotspots and high-risk groups only^3^. However, this approach has been questioned by some medical practitioners^4^, which reckon that Malaysia should conduct COVID-19 testings as wide as possible (i.e., mass testing) so that these tests would be able to pick up most (if not all) of the possible cases; this crucial information can help government to devise an effective contact tracing strategy, and these possible patients could be placed on 14-day quarantine to contain the spread of the disease.

Motivated by the aforementioned problem, we utilise a simple epidemiological system with limited medical resources component, which results in the possibility of under-testing issue. Inspired by the previous reports^5, 6^, the problem of false detection (i.e., false negative tests) and the possibility of ‘reinfection’ are also investigated by employing our Susceptible-Infected-Removed-Susceptible (SIRS)-type system, based on daily reported cases in Malaysia, which is used a case study.

## 2. MODEL AND METHODS

To model the combined effects of limited medical resources and false detection problems in shaping the COVID19 pandemic, we revisit a simple ordinary differential equation (ODE) model of SIRS-type^7–10^:

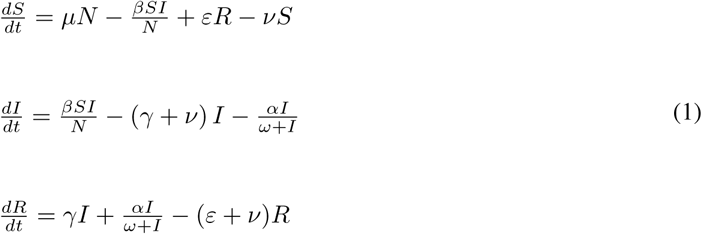

where population of *N* individuals are divided into three compartments: *S*(*t*) corresponds to susceptible population, which can be infected (with transmission rate of *β*); *I*(*t*) corresponds to the number of infectious individuals and *R*(*t*) corresponds to the removal class where those individuals who have recovered (with recovery rate of *γ*) or dead are removed from the population. The parameters *µ* and *ν* correspond to the birth and death rates of the population, respectively. To investigate the impacts of false detection on COVID-19 transmission dynamics, we allow for some proportions of individuals from the removal compartment to re-enter the susceptible class (at a transfer rate of *ε*); this component is also used as a proxy to account for the likelihood of ‘reinfection’: it has been reported that, until this research work is conducted, we do not know whether some of the COVID-19 patients who have recovered are truly been ‘reinfected’, or, at the time of their ‘recovery’, they still had low levels of the virus in their systems^5^. There is also no evidence whether people have herd or long-term immunity against this new coronavirus attack^6^. Another concern in controlling this pandemic is about the issue of under-testing for COVID-19, which is evident in some developing countries; this situation occurs due to limited medical resources such as tests, drugs and hospital beds^3, 11, 12^. To study the consequence of the limited medical resources on the spread of COVID-19 outbreak, we employ the term of 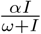, which is motivated by Zhou and Fan^9^: the parameter 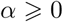 represents the medical resources supplied per unit time and *ω >* 0 corresponds to half-saturation constant, which measures the efficiency of the medical resource supply (i.e., if *ω* is smaller, then the efficiency is higher). In fact, this quantity also represents the efficiency of the supply of available medical resources, and this situation would also depend on other factors such as the control strategies (e.g., quarantine, movement control order) and the production of drugs, etc. All of the parameters are positive in this model to depict the biologically-meaningful phenomenon of transmission dynamics.

The model (1) has two types of steady states, which are disease-free and pandemic equilibria: (i) disease-free equilibrium,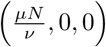; (ii) pandemic equilibrium, (*S*^∗^,*I*^∗^,*R*^∗^). Local stability analysis is employed so as to determine stability of each of these steady states. To obtain basic reproduction numbers, *R*_0_, we used the next generation matrix method (and the correstness of these quantities are checked using eigenvalues analysis). The simplest SIR system can obtained from the model (1) by letting *µ* = *ν* = *ε* = *α* = 0, with basic reproduction number, *R*_0_*S*, is as follows:

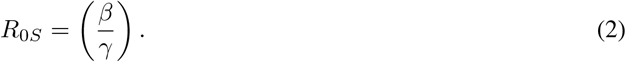

When the possibility of limited medical resources is relaxed (i.e., *α* = 0 and other parameters are positive), basic reproduction number, *R*_0_*_u_*, is given by:

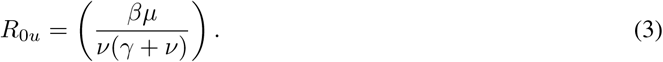

Considering the case of a limited medical resources (i.e., *α* > 0), this situation leads to different threshold of basic reproduction number, *R*_0_*_L_*:

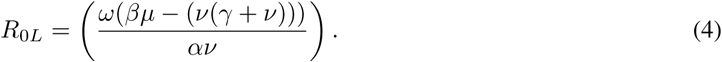

Numerical simulations are conducted for the model (1) and the results are discussed in the next sections. As a case study, the transmission dynamics are investigated using daily confirmed cases in Malaysia, where some of the parameters are estimated based on the fitting of the model (1) to COVID-19 cases in the interval between March 24 and April 23, 2020^13^. Unless otherwise stated, parameter values used in the simulation are given in figure captions. In all cases, we employed numerical simulation using MATLAB `ode15s` solver. We also used numerical continuation package XPPAUT to perform one-and two-parameter bifurcation analysis; the stable and unstable steady states are tracked as a model parameter changes.

## 3. RESULTS AND DISCUSSIONS

First, we consider a simple scenario shown by Fig. 1A where the assumption of limited medical resources is relaxed (i.e., *α* = 0) and there is no possibility of false detection or ‘reinfection’ (i.e., *ε* = 0) in this epidemiological system. This situation corresponds to the dynamics of basic SIR model (solid lines) with *R*_0_*_S_* = 2.7586: we observe that the active cases increase on daily basis until the infection curve (solid red) peaks around first week of April; this observation is in agreement with the number of reported active COVID19 cases in Malaysia (cyan asterick), which peaks around April 5–8, 2020 with the highest 2596 active cases recorded on April 5. Consequently, the number of active cases start to decrease and SIR model predicts that infection will be excluded in the long run.

**Fig. 1.**
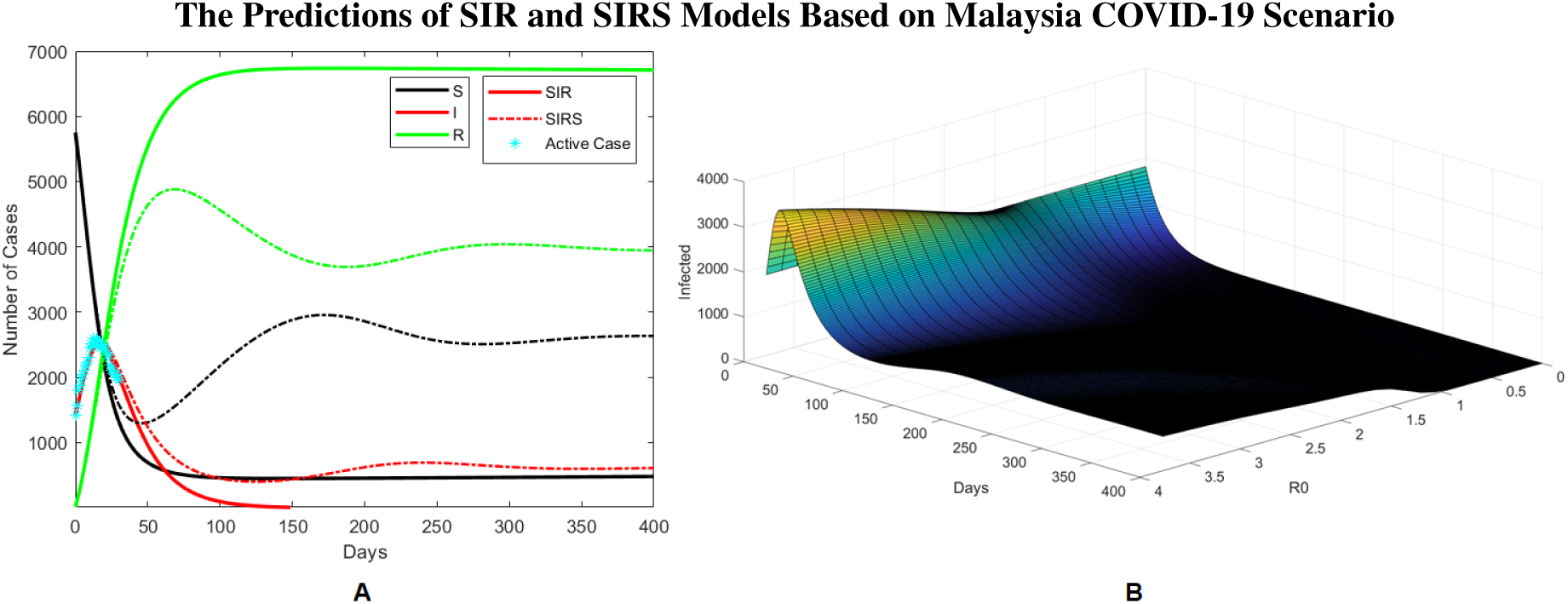
Numerical simulation results of SIR (*ε* = *α* = 0) and SIRS (*ε* > 0, *α* = 0) models with the assumption of limited medical resources is relaxed. (A) Time series results of SIR (solid lines) and SIRS (dashed lines) models and comparison with the number of active cases in Malaysia (cyan asterick). (B) The daily number of active cases in Malaysia as *R*_0_*_u_* varies based on predictions of SIRS model: as *R*_0_*_u_* < 1, the pandemic can be excluded; when *R*_0_*_u_* > 1, the pandemic persists for a long period of time. Other parameter values: *β* = 0.16 (fitted), *γ* = 0.058 (fitted), *µ* = *ν* = 0.00002^14^, *N* = *S* + *I* + *R* and *ε* = 0.0009 (fitted).

The predictions of SIR represents an ‘ideal’ situation, assumming medical resources to fight this pandemic are being allocated enough, apart from citizens have been screeneed massively and detected (i.e., quarantined) quickly; additionally, it is also assumed that the test results are reliable and can be trusted. In reality, some nations especially the developing countries (e.g., Malaysia, Vietnam, etc.), face the risk of under-screening due to limited medical resources, and also the possibility of false detection. To illustrate the impacts of inaccurate detection or ‘reinfection’ problem using SIRS system (1), we let *ε* > 0 (keeping *α* = 0) and the results are plotted in Fig. 1A (dashed lines). In comparison to our previous predictions of COVID-19 transmission dynamics in Malaysia, we realise that SIR and SIRS observations are indistinguishable before the infection peak; after the infected trajectories reach the peak point, SIRS results show worst-case scenario with increasing number of infectious (dashed red) and susceptible (dashed black) individuals; in this case, SIRS demonstrates that the pandemic will persist for a long time. This is in agreement with our analysis of basic reproduction number: *R*_0_*_u_* is calculated to be 2.7577 using equation (3) for Malaysia’s COVID-19 scenario (in practice, *R*_0_*_u_* > 1 implies a pandemic outbreak). In order to control this pandemic, it is vital to employ proper mitigation measures such as quarantine, travel restrictions and social distancing; these strategies can help to push *R*_0_*_u_* to be less than 1, which in principle can eradicate this pandemic in a long run (Fig. 1B).

**Fig. 2.**
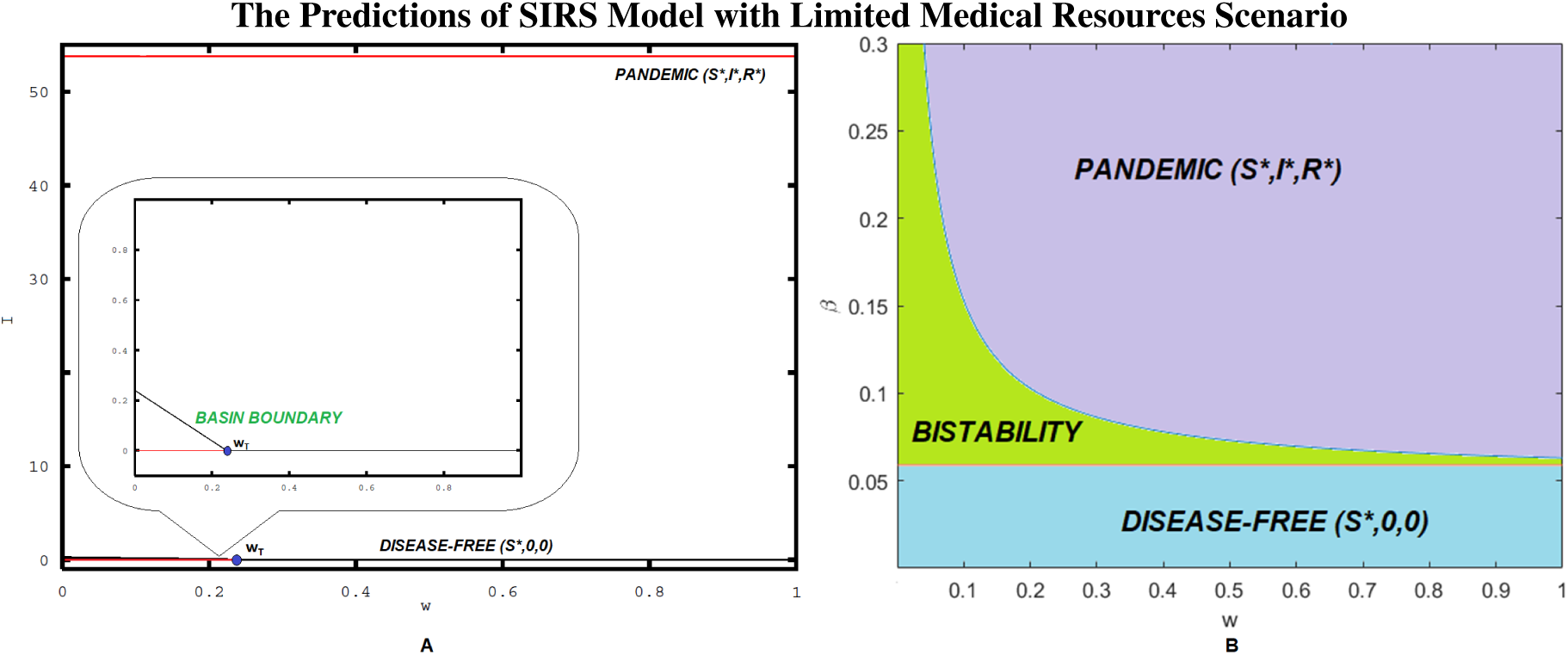
Bifurcation analysis results of the model (1) with limited medical resources and possibility of false detection. (A) A one-parameter bifurcation diagram when *β* = 0.095 as *ω* varies with threshold *ω_T_* respresents transcritical bifurcation (blue point). (B) A two-parameter bifurcation diagram in (*ω*, *β*)-plane with different colour regions represent transmission dynamics of the system. Other parameter values: *µ* = *ν* = 0.00002^14^, *N* = *S* + *I* + *R*, *ε* = 0.0009 (fitted), *γ* = 0.053 (fitted) and *α* = 0.01^9^.

However, the use of basic reproduction number should be interpreted with care in managing the deadly and fast evolving pandemic outbreaks such as COVID-19. Other counterintuitive observations are also possible even when *R*_0_ < 1. To illustrate this observation, we consider our SIRS model with limited medical resources scenario (i.e., *ε* > 0 and *α* > 0). Fig. 2A depicts an example of bifurcation analysis of the model (1) with *β* = 0.095 and *ε* = 0.0009 as the efficiency of the medical resource supply (*ω*) is varied. There occurs one critical value i.e., *ω_T_*, which corresponds to transcritical bifurcation (blue point). From an epidemiological viewpoint, *ω_T_* is a minimum threshold of medical resource supply to ensure the infection dynamics can be reduced efficiently and effectively. There are also several branches of steady states: (i) the upper branch (pandemic equilibrium) is always stable (red curve); (ii) lower branch (disease-free equilibrium) can be stable (red curve) or unstable (black curve), depending on the values of *ω*. When *ω < ω_T_*, we observe the occurrence of alternative stable states, with both pandemic and disease-free equilibria are stable; convergence to either one of these steady states depends on initial abundances of individuals in susceptible, infected and removal classes. When *ω>ω_T_*, this situation leads to rapidly evolving pandemic outbreaks. Closer examination of these outcomes reveals that *R*_0_*_L_* of SIRS model (1) with limited medical resources scenario is 0.8396 (calculated using equation (4)) when *ω* < *ω_T_*. While basic reproduction number is less than 1 in this case, it is observed that the COVID-19 pandemic is still likely to occur due to a relatively small basin of attractions of disease-free equilbrium (lower branch steady state), which can cause a catastrophic shift to pandemic equilbrium (upper branch steady state). In general, the two bistable attractors are separated by a basin boundary (unstable pandemic equilibrium), which determines the long-term dynamics of this epidemiological system.

Based on equation (4), we also notice some mechanisms that can result in *R*_0_*_L_* to be shifted to larger values: (i) higher magnitudes of *ω* indicate inefficiency or lack of medical resource supply, which consequently could lead to pandemic outbreaks; (ii) as the transmission rate, *β*, is higher, the pace of pandemic spread increases significantly. To demonstrate these mechanisms, we plotted a two-parameter bifurcation diagram in Fig. 2B as *ω* and *β* are varied. The blue and red curves correspond to transcritical bifurcations, and the interaction between these two local bifurcations determine the dynamical behaviours of this epidemiological system and separate the parameter space into three different regions: (i) purple region corresponds to pandemic equilibrium; (ii) cyan region corresponds to disease-free equilibrium; (iii) green region corresponds the appearance of alternative stable states between pandemic and disease-free outcomes. We can see that an increase in *ω* and *β* will dramatically increase *R*_0_*_L_*, and this situation can be detrimental in controlling the negative impacts of COVID19, as more outcomes of pandemic outbreak are possible.

## 4. CONCLUSION

In this study, the most important case that we identified is the possibility of a COVID-19 pandemic through bistable behaviour, even when the basic reproduction number is less than unity. This scenario is observed when certain complexities such as limited medical resources and false detection problems are incorporated into the model, which make the epidemiological system to be more realistic. Based on these findings, we caution policy makers not to make their decisions solely based on the guidance of the basic reproduction number only, which clearly courting trouble. This is because other factors such as alternative stable states phenomenon can interfere with distinct epidemiological forces and lead to catastrophic shifts in transmission dynamics. We recommend for the existing policies in Malaysia e.g., movement control order, quarantine and social distancing to be further practiced so as to break the chain of infections and flatten the infection curve, given the country’s limited medical resources scenario and other pressing issues such as the risk of under-screening. Careful consideration of these issues will minimise spurious conclusions and maximise the amount of knowledge extracted from our modelling practices, which can also promote an evidence-based decision making in public health sector.

## Data Availability

Not related

## Acknowledgement

This research is supported by the Universiti Sains Malaysia (USM) Fundamental Research Grant Scheme (FRGS) No. 203/PMATHS/6711645. The authors would like to thank Dr. Kamarul Imran Musa and Dr. Wan Nor Arifin for many fruitful discussions on the situation of COVID-19 pandemic in Malaysia.

## REFERENCES

1. Huang Y, Yang L, Dai H, Tian F, Chen K (2020) Epidemic situation and forecasting of COVID-19 in and outside China. Bull World Health Organ.

2. Li R, Pei S, Chen B, Song Y, Zhang T, Yang W, Shaman J (2020) Substantial undocumented infection facilitates the rapid dissemination of novel coronavirus (SARS-COV 2). Science.

3. Annuar A (2020) Targeted testing better use of resources in covid-19 fight, says Health DG. Malay Mail. [https://www.malaymail.com/news/malaysia/2020/04/13/targeted-testing-better-use-of-resources-in-covid-19-fight-says-health-d-g/1856325]

4. Nordin MM, Ismail Z (2020) Malaysia must ramp up testing. The Star. [https://www.malaymail.com/news/malaysia/2020/04/13/targeted-testing-better-use-of-resources-in-covid-19-fight-says-health-d-g/1856325]

5. Malkov E (2020) Covid-19 in the us: Estimates of scenarios with possibility of reinfection. *Available at SSRN 3569097*.

6. Shi Y, Wang Y, Shao C, Huang J, Gan J, Huang X, Bucci E, Piacentini M, et al (2020), Covid-19 infection: the perspectives on immune responses. Cell Death & Differentiation.

7. Kermack WO, McKendrick AG (1927) A contribution to the mathematical theory of epidemics. Proceedings of the Royal Society of London Series A 115, 700–721.

8. Hethcote HW (2000) The mathematics of infectious diseases. SIAM Review 42, 599–653.

9. Zhou L, Fan M (2012) Dynamics of an SIR epidemic model with limited medical resources revisited. Nonlinear Analysis Real World Applications 13, 312–324.

10. Muroya Y, Li H, Kuniya T (2014) Complete global analysis of an SIRS epidemic model with graded cure and incomplete recovery rates. Journal of Mathematical Analysis and Applications 410, 719–732.

11. Rokx C, Schieber G, Harimurti P, Tandon A, Somanathan A (2004) Health Financing in Indonesia. The World Bank.

12. Hermawan A (2020) With limited testing capability, Indonesia may battle COVID19 blindfolded. The Jakarta Post. [https://www.thejakartapost.com/academia/2020/04/18/with-limited-testing-capability-indonesia-may-battle-covid-19-blindfolded.html]

13. MOH (2020) Press statement updates on COVID-19 situation in Malaysia. Ministry of Health Malaysia.

14. Mummert A, Otunuga OM (2019) Parameter identification for a stochastic seirs epidemic model: case study influenza. Journal of Mathematical Biology 79, 705–729.

